# Healthcare utilisation in people with long COVID: an OpenSAFELY cohort study

**DOI:** 10.1101/2023.12.21.23300305

**Authors:** Liang-Yu Lin, Alasdair D Henderson, Oliver Carlile, Iain Dillingham, Ben FC Butler-Cole, Michael Marks, Andrew Briggs, Mark Jit, Laurie A Tomlinson, Chris Bates, John Parry, Sebastian CJ Bacon, Ben Goldacre, Amir Mehrkar, Brian MacKenna, The OpenSAFELY Collaborative, Rosalind M Eggo, Emily Herrett

## Abstract

**Background:** Long COVID, characterised by various symptoms and complications, potentially increases healthcare utilisation and costs. However, its impact on the NHS remains to be determined.

**Objective:** This study aims to assess the healthcare utilisation of individuals with long COVID.

**Methods:** With the approval of NHS England, we conducted a matched cohort study using primary and secondary care data via OpenSAFELY, a platform for analysing anonymous electronic health records. The long COVID exposure group, defined by diagnostic codes, was matched with five comparators without long COVID between Nov 2020 and Jan 2023. We compared their total healthcare utilisation from GP consultations, prescriptions, hospital admissions, A&E visits, and outpatient appointments. Healthcare utilisation and costs were evaluated using a two-part model adjusting for covariates. Using a difference-in-difference model, we also compared healthcare utilisation after long COVID with pre-pandemic records.

**Results:** We identified 52,988 individuals with a long COVID diagnosis, matched to 264,867 comparators without a diagnosis. In the 12 months post-diagnosis, there was strong evidence that those with long COVID were more likely to use healthcare resources (OR: 8.07, 95% CI: 7.54 – 8.64), and have 49% more healthcare utilisation (RR: 1.49, 95% CI: 1.47 – 1.50). Our model estimated that the long COVID group had 30 healthcare visits per year (predicted mean: 29.23, 95% CI: 28.58 - 29.92), compared to 16 in the comparator group (predicted mean visits: 16.04, 95% CI: 15.73 - 16.36). Individuals with long COVID were more likely to have non-zero healthcare expenditures (OR = 7.47, 95% CI = 7.02 – 7.95), with costs being 43% higher than the comparator group (cost ratio = 1.43, 95% CI: 1.38 – 1.49). The long COVID group costs approximately £2,500 per person per year (predicted mean cost: £2,562.50, 95% CI: £2,335.60 - £2,819.22), and the comparator group costs £1,500 (predicted mean cost: £1,527.43, 95% CI: £1,404.33 - 1,664.45.) Historically, individuals with long COVID utilised healthcare resources more frequently, but their average healthcare utilisation increased more after being diagnosed with long COVID, compared to the comparator group.

**Conclusion:** Long COVID increases healthcare utilisation and costs. Public health policies should allocate more resources towards preventing, treating, and supporting individuals with long COVID.

## Introduction

After infection with SARS-CoV-2, symptoms usually resolve in four weeks; however, for some people the symptoms persist. The National Institute for Health and Care Excellence (NICE) defines symptoms lasting from four to 12 weeks as “ongoing symptomatic COVID-19” and longer than 12 weeks as “post-COVID-19 syndrome.” According to the NICE guidelines, ongoing symptomatic COVID-19 and post-COVID-19 syndrome both refer to long COVID. Common symptoms of long COVID include weakness, general malaise, fatigue, concentration impairment (known as “brain fog”), and breathlessness (1). In March 2023, the Office for National Statistics reported that about 1.9 million people, (approximately 2.9% of the UK population), had long COVID symptoms (2).

The persistent symptoms of long COVID affect quality-of-life (3), and patients seek care for their symptoms (4–6). Evidence from the UK and other countries has demonstrated an increase in healthcare use and costs in groups with long COVID (7–9). However, many of these studies define long COVID based on COVID-19 testing, which introduces selection bias due to testing policy, and reporting of testing (10–12).

There is an urgent need to fully quantify the healthcare use of patients with long COVID, to allow healthcare planning decisions and to properly quantify the impact of the COVID-19 pandemic on the healthcare system. Therefore, our study aims to investigate the healthcare utilisation of people with long COVID, factors associated with increased utilisation, and the associated cost to the NHS.

## Methods

### Data Source

All data were linked, stored and analysed securely within the OpenSAFELY platform https://opensafely.org/. Data include pseudonymized data such as coded diagnoses, medications and physiological parameters. No free text data are included. All code is shared openly for review and re-use under the MIT open license (https://github.com/opensafely/openprompt_health_utilisation). Detailed pseudonymised patient data is potentially re-identifiable and therefore not shared. We rapidly delivered the OpenSAFELY data analysis platform without prior funding to deliver timely analyses on urgent research questions in the context of the global COVID-19 health emergency: now that the platform is established we are developing a formal process for external users to request access in collaboration with NHS England; details of this process are available at OpenSAFELY.org. Primary care records managed by the GP software provider, TPP, were linked to ONS death data, emergency care attendance, hospital admission, outpatient clinic visit records and costs data through OpenSAFELY.

### Study population and eligibility

We conducted a matched retrospective cohort study using electronic health records and performed two comparisons: one contemporary and one historical (**Figure 1**). A ‘contemporary’ comparison was designed to demonstrate differences in healthcare utilisation after long COVID diagnosis at a given calendar time compared to matched controls, when the pandemic or seasonal variation in illness might affect utilisation across the whole population. A ‘historical’ comparison was designed to understand differences between those with and without long COVID in terms of their previous healthcare utilisation and to examine the change in use within and between groups over time.

**Figure 1.**
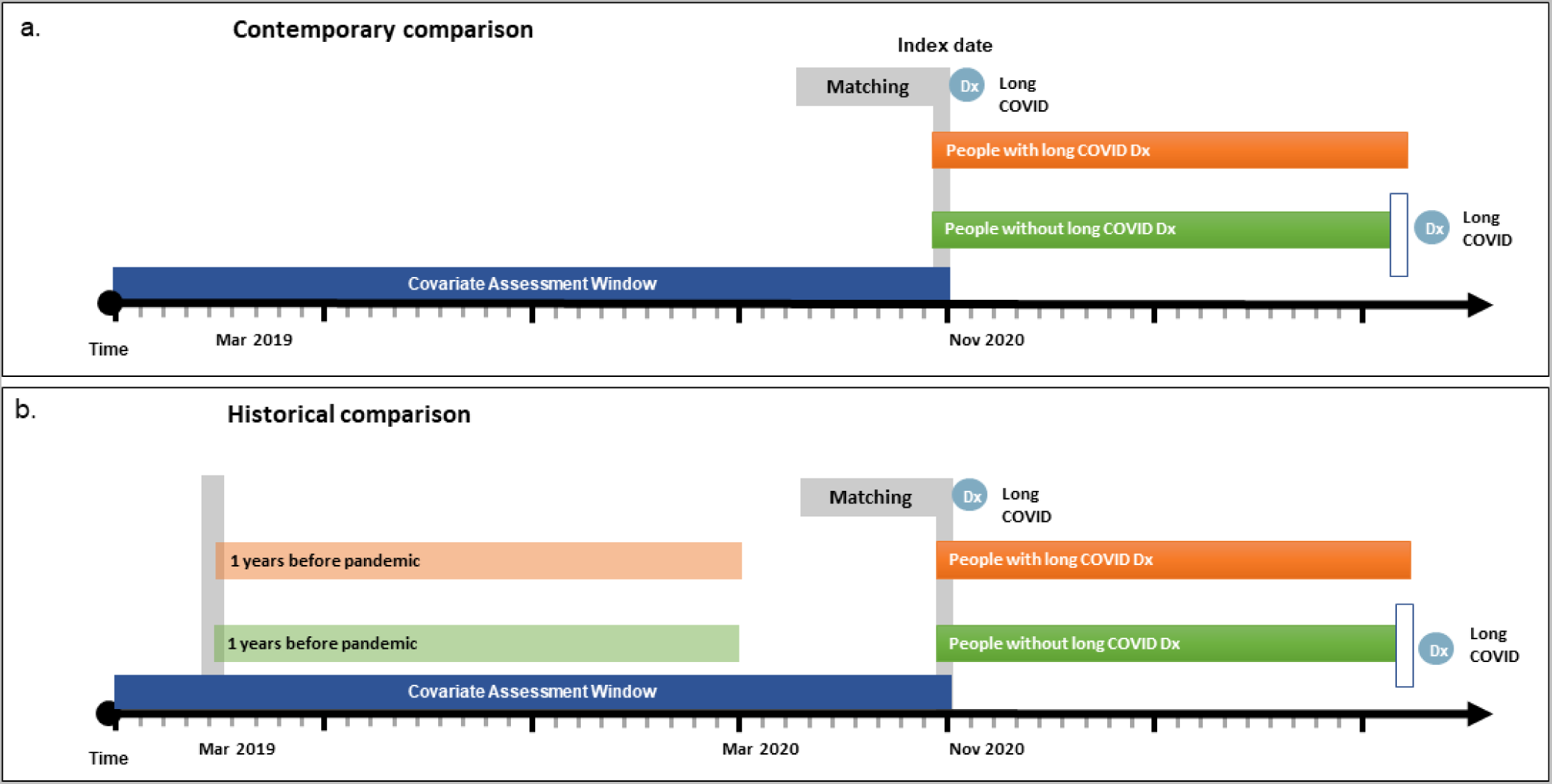
The study design. a) Contemporary comparisons; b) Historical comparisons

In the contemporary comparison, we matched each patient by age, sex and region with a recorded long COVID diagnosis to five controls without, using long COVID diagnosis as the index date. For the comparator group, the index date was assigned when they were matched to a long COVID patient. We followed the cohort from index date to the earliest of: 1. date of death; 2. end of GP registration; 3. receipt of a resolved long COVID SNOMED code (1326351000000108) among the exposed group; 4. receipt of a long COVID diagnosis among the unexposed group; 5. 31st January 2023. We compared their healthcare utilisation in the 12 months after index date (**Figure 1a**). We included adults aged 18 or over who had been registered with a GP practice using TPP software for at least three months prior to 1st November 2020. To exclude bias due to unusual coding practices, we excluded patients registered with GP practices that did not use at least one long COVID diagnostic code between November 2020 and January 2023.

In the historical comparison, we took matched sets of patients from the contemporary comparison, who were additionally registered with their GP between March 2019 and the index date. Among the matched sets we examined (i) their historical healthcare utilisation between March 2019 and March 2020 and (ii) their contemporary utilisation after the index date (**Figure 1b**). The difference in utilisation between exposed (long COVID) patients and unexposed patients was assessed using a difference-in-difference analysis.

### Exposures

The primary exposure of interest was long COVID, defined by SNOMED-CT codes recorded in primary care, (including diagnostic, referral, and long COVID assessment codes (13) (**Supplementary Table 1**)). The date of the first long COVID code in the primary care records was defined as the index date.

### Outcomes

For the contemporary comparison, the primary outcome was total healthcare resource utilisation in the 12 months following the first record of long COVID in primary care. Total healthcare resource utilisation was calculated by combining: 1. primary care utilisation, including consultation with a GP and/or prescription of medications; 2. all-cause accident and emergency (A&E) visits, defined using A&E arrival records; 3. all-cause hospital outpatient visits, defined as admission to hospitals for more than one day; and 4. all-cause hospital outpatient clinic visits. For each type of healthcare utilisation, multiple healthcare visits on the same date were regarded as one visit.

The secondary outcome was the cumulative total healthcare costs in the 12 months after the index date. This was calculated by combining: 1. primary care costs, including GP consultations and prescriptions; 2. hospitalisation costs; 3. A&E costs, and 4. outpatient clinic costs. We estimated the cost of a GP consultation by multiplying the GP visit counts and the average cost for a GP consultation in 2021/2022 (£41) (14). To estimate the cost of GP prescriptions, we multiplied the frequency of prescriptions for each BNF chapter by the average cost of medications in that chapter in 2021/2022(12). The hospital admission costs, A&E costs, and hospital outpatient costs were provided by NHS England (15). We additionally analysed the four components of utilisation and cost separately.

For the historical comparison, the outcome was the difference between total healthcare resource utilisations (as defined above) before (i.e. during the period March 2019-March 2020) and after being diagnosed with long COVID (12 months after index date).

### Covariates

In our analyses, we determined covariates by using a DAG (**Supplementary Figure 1**), including age, sex, ethnicity, region and Index of Multiple Deprivation (IMD) quintile. We also included underlying chronic diseases, which included asthma, obesity/overweight, previous psychiatric conditions, and the level of multi-morbidity. The level of multi-morbidity was defined by categorising the number of chronic diseases listed in **Supplementary Table 2**. The covariate assessment period was five years before November 2020. We also considered previous hospital admissions due to COVID-19 and the number of COVID-19 vaccination doses (any vaccine) received before the index date.

### Statistical analysis

We first compared the distribution of demographic factors, underlying comorbidities, and socioeconomic factors in the long COVID and comparator groups. Categorical variables were assessed using Chi-square statistics, and the mean and standard deviation of continuous variables were compared using a t-test. In the contemporary comparison, because the distribution of healthcare visits and costs were zero-inflated and right-skewed (**Supplementary Figure 2a and 2b**), we implemented a two-part model to analyse the healthcare utilisation and cost data (16,17). In brief, the first part of the model is a binomial model, estimating the probability of non-zero healthcare visits or non-zero healthcare costs; the second part of the model is a truncated GLM model conditioning on people with non-zero healthcare visits and non-zero healthcare costs. In the second part of the analysis, we used a negative binomial model to estimate the overall healthcare utilisation rate ratio and a Gamma GLM model to assess the total healthcare cost ratio. We examined the over-dispersion of the data by running a Poisson regression and examining the ratio between residual deviance and the degree of freedom. If the ratio was greater than 1, we applied a negative binomial model in the second part of our model and carried out a Poisson regression model if the ratio was close to 1. We further applied a prediction function to the regression model outputs, multiplying the probability of non-zero healthcare visits and the predicted healthcare visits, to obtain the predicted average healthcare utilisation and costs on the absolute scale.

For the historical comparison, we conducted a difference-in-difference (DID) analysis evaluating the change in healthcare utilisations before the pandemic compared to after a long COVID diagnosis. We created a time variable and categorised healthcare visits between March 2019 and March 2020 as “historical records” (pre-pandemic) and healthcare visits after the index date as “contemporary records.” By fitting this time variable interacting with the exposure variable in the two-part model, we could compare the healthcare utilisation difference before and after the index date within the exposure and the comparator groups, and then further calculate the difference between these two values. Similar to the contemporary comparison, we also used a prediction function to multiply the probability of non-zero healthcare visits and the predicted healthcare visits, to estimate the average healthcare visits before and after long COVID diagnoses on an absolute scale. The common trend assumption of DID was examined by comparing the average healthcare utilisation in the exposure and comparator groups over time (**Supplementary Figure 4.**)

### Sensitivity analysis

We carried out a series of sensitivity analyses. First, some patients in our study had healthcare visit records but the associated healthcare cost data were missing. We imputed missing secondary cost data by estimating the mean cost for one visit, appointment, or admission episode from people with both healthcare cost and healthcare visit records. Second, we stratified our analyses by sex, age group and previous hospital admission due to COVID-19. The stratum-specific results were obtained by fitting an interaction term between exposure and the stratifying variables. The interaction was examined using a likelihood ratio test. Third, because people with outcomes can only be identified if they visited a healthcare provider, we restricted the main analyses to people who had ever consulted a GP 1 year before the 1st of November 2020. Fourth, to balance the chance of getting long COVID between groups, we restricted the analyses to people who had tested positive for COVID.

### Software and reproducibility

Data management was performed using Python 3.8, with analysis carried out using R 4.0. Code for data management and analysis as well as codelists archived online (https://github.com/opensafely/openprompt_health_utilisation). All iterations of the pre-specified study protocol are archived with version control (https://github.com/opensafely/openprompt_health_utilisation/blob/cd8ecce1e12018756375013cd3b27a25880d85a4/OpenPROMPT_longCOVID_healthcare_utilisation_protocol.pdf). We report our results following the RECORD reporting guideline (18) (**Supplementary Table 3**).

### Information governance and ethical approval

NHS England is the data controller of the NHS England OpenSAFELY COVID-19 Service; TPP is the data processor; all study authors using OpenSAFELY have the approval of NHS England (19). This implementation of OpenSAFELY is hosted within the TPP environment which is accredited to the ISO 27001 information security standard and is NHS IG Toolkit compliant (20). This study was conducted as part of the “Quality-of-life in patients with long COVID: harnessing the scale of big data to quantify the health and economic costs study (OpenPROMPT)”, which was approved by and the LSHTM Research Ethics Committee (ref 28030).

### Patient and Public Involvement and Engagement (PPIE)

In the OpenPROMPT research group, we have three representatives from the public through our Patient and Public Involvement and Engagement (PPIE) initiative. These representatives attended progress meetings every six months to provide feedback and insights on our work. Furthermore, we had two online open workshops inviting individuals living with long COVID, aiming to better understand their lived experiences and healthcare-seeking behaviours. In addition, OpenSAFELY has developed a publicly available website https://www.opensafely.org/, through which they invite any patient or member of the public to make contact regarding the broader OpenSAFELY project.

## Results

### Study population

We identified 52,988 people with long COVID and 164,872 matched comparators (**Table 1**). There were more females than males, and people aged 40 to 59 comprised 50% of the sample. The long COVID group had a higher proportion of white ethnicity, obesity, asthma, mental health diseases, and other comorbidities, compared to the comparator group. They were also more likely to have been hospitalised for COVID, and had received more COVID vaccines. Less than 40% of people in both groups had a linked positive SARS-CoV-2 test before the index date (**Table 1**). There were more missing values for IMD quintile (1.8%), ethnicity (14.7%) and BMI categories (8.3%) (**Supplementary Table 4**). Additionally, admission cost, A&E visit cost, and outpatient clinic cost data were only available among 2 to 9% of study population (**Supplementary Table 5**).

**Table 1.** Distribution of demographic factors.

| Factors | Level | Total (N, %) | Long covid exposure (N, %) | Comparator (N, %) | P-value |
| --- | --- | --- | --- | --- | --- |
| Sex | Female | 317,852 (100.0) | 33,804 (63.8) | 168,981 (63.8) | 0.99 |
|  | Male |  | 19,184 (36.2) | 95,883 (36.2) |  |
| Age | Mean (SD) | 317,852 (100.0) | 48.0 (14.4) | 48.0 (14.4) | 0.98 |
| Age categories | 18-29 | 317,855 (100.0) | 6,638 (12.5) | 33,188 (12.5) | 1.00 |
|  | 30-39 |  | 10,050 (19.0) | 50,234 (19.0) |  |
|  | 40-49 |  | 13,241 (25.0) | 66,194 (25.0) |  |
|  | 50-59 |  | 13,236 (25.0) | 66,152 (25.0) |  |
|  | 60-69 |  | 6,199 (11.7) | 30,985 (11.7) |  |
|  | 70+ |  | 3,624 (6.8) | 18,111 (6.8) |  |
| Ethnicity | White | 271,276 (85.3) | 40,984 (89.0) | 195,323 (86.7) | <0.01 |
|  | Mixed |  | 539 (1.2) | 2,800 (1.2) |  |
|  | South Asian |  | 3,052 (6.6) | 16,226 (7.2) |  |
|  | Black |  | 832 (1.8) | 5,624 (2.5) |  |
|  | Other |  | 655 (1.4) | 5,241 (2.3) |  |
| BMI categories | Normal weight | 291,410 (91.7) | 851 (1.7) | 5,330 (2.2) | <0.001 |
|  | Underweight |  | 13,320 (26.4) | 79,908 (33.2) |  |
|  | Overweight |  | 15,792 (31.3) | 78,912 (32.7) |  |
|  | Obese |  | 20,450 (40.6) | 76,847 (31.9) |  |
| Index of multiple deprivation (IMD) | least deprived | 312,032 (98.2) | 10,457 (20.1) | 52,359 (20.1) | 0.93 |
|  | 2<sup>nd</sup> deprived |  | 10,445 (20.1) | 52,340 (20.1) |  |
|  | 3<sup>rd</sup> deprived |  | 10,479 (20.2) | 51,976 (20.0) |  |
|  | 4<sup>th</sup> deprived |  | 10,349 (19.9) | 52,020 (20.0) |  |
|  | Most deprived |  | 10,272 (19.8) | 51,332 (19.7) |  |
| Region | East | 317,837<br>(100.0) | 10,162 (19.2) | 50,807 (19.2) | 1.00 |
|  | East Midlands |  | 7,513 (14.2) | 37,559 (14.2) |  |
|  | London |  | 2,400 (4.5) | 11,995 (4.5) |  |
|  | North East |  | 4,161 (7.9) | 20,805 (7.9) |  |
|  | North West |  | 6,068 (11.5) | 30,340 (11.5) |  |
|  | South East |  | 3,707 (7.0) | 18,535 (7.0) |  |
|  | South West |  | 8,267 (15.6) | 41,353 (15.6) |  |
|  | West Midlands |  | 1,675 (3.2) | 8,374 (3.2) |  |
|  | Yorkshire and The Humber |  | 9,020 (17.0) | 45,096 (17.0) |  |
| Asthma | No asthma | 317,852<br>(100.0) | 44,248 (83.5) | 239,545 (90.4) | <0.01 |
|  | Have asthma |  | 8,740 (16.5) | 25,319 (9.6) |  |
| Mental health issues | No mental health issues | 317,852<br>(100.0) | 45,514 (85.9) | 241,066 (91.0) | <0.01 |
|  | Have mental health issues |  | 7,474 (14.1) | 23,798 (9.0) |  |
| Number of comorbidities | 0 | 317,852<br>(100.0) | 47,590 (89.8) | 243,891 (92.1) | <0.01 |
|  | 1 |  | 4,822 (9.1) | 19,196 (7.2) |  |
|  | 2 |  | 513 (1.0) | 1,644 (0.6) |  |
|  | 3 or more |  | 63 (0.1) | 133 (0.1) |  |
| Previous hospitalisation due to COVID | No | 317,852<br>(100.0) | 48,333 (91.2) | 263,159 (99.4) | <0.01 |
|  | Yes |  | 4,655 (8.8) | 1,705 (0.6) |  |
| Number of COVID vaccine received at index date | 0 dose | 317,852<br>(100.0) | 8,454 (16.0) | 54,697 (20.7) | <0.01 |
|  | 1 dose |  | 5,216 (9.8) | 23,770 (9.0) |  |
|  | 2 doses |  | 17,259 (32.6) | 80,898 (30.5) |  |
|  | 3 or more doses |  | 22,059 (41.6) | 105,499 (39.8) |  |
| Had long COVID diagnoses | No | 317,852<br>(100.0) | 0 (0.0) | 264,267 (99.8) | <0.01 |
|  | Yes |  | 52,988<br>(100.0) | 597 (0.2) |  |
| Had been tested positive for COVID-19 | No | 317,852<br>(100.0) | 32,005 (60.4) | 182,442 (68.9) | <0.01 |
|  | Yes |  | 20,983 (39.6) | 82,422 (31.1) |  |

### Healthcare utilisation and cost among people with long COVID

After adjusting for covariates, there was strong evidence that people with long COVID were more likely to use healthcare resources (**Fig 2a**, first part: odds ratio (OR): 8.29, 95% CI: 7.74 – 8.87), and further among those who visited their healthcare providers, there was strong evidence that people with long COVID had a higher rate of total healthcare utilisation (**Fig 2a**, second part: rate ratio (RR): 1.49, 95% CI: 1.48 – 1.51). The predicted model shows that on average, the long COVID group had nearly 30 healthcare visits per year (predicted mean: 29.23, 95% CI: 28.58 - 29.92), while the comparator group had 16 visits per year (predicted mean visits: 16.04, 95% CI: 15.73 - 16.36) (**Fig 2**, average total healthcare utilisations). The increase in healthcare utilisation persisted across all healthcare types (Figure 2b-f) and in those using any healthcare services, the rate ratio was highest for GPs, and lowest for inpatient hospital stays.

**Figure 2.**
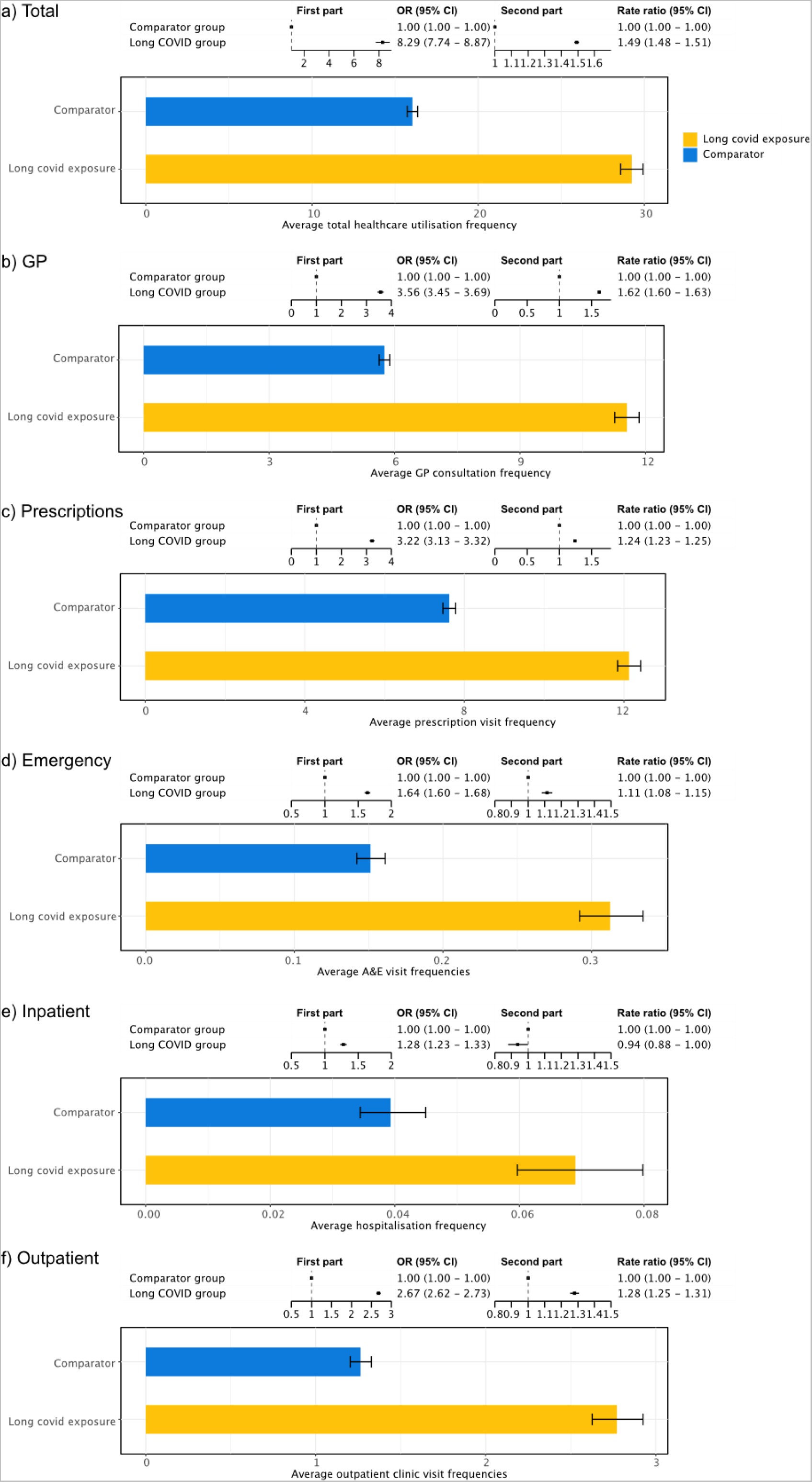
Healthcare utilisation among people with long COVID. Each subfigure shows the first and second part of the model in the upper panel, where the first part gives the odds ratio (OR) of healthcare resource utilisation and the second part is the rate ratio (RR) of healthcare utilisation between people with long COVID and comparator groups, conditioned on people having used healthcare resources. Each lower panel shows the predicted average healthcare utilisation in long COVID and Comparator groups. Values are shown for a) Total healthcare utilisation, and then separately for each part of the total: b) GP visits, c) prescriptions, d) Emergency care at A&E, e) Inpatient hospitalisations, f) Outpatient visits.

We also estimated healthcare costs; after adjusting for covariates, there was strong evidence that people with long COVID were seven times more likely to have non-zero healthcare costs (**Figure 3a**, first part, OR = 7.66, 95% CI = 7.20 – 8.15). Among people with non-zero healthcare costs, the total costs for people with long COVID were 44% higher than those of the comparator groups (**Figure 3a**, second part, OR =1.44, 95% CI: 1.39 – 1.50). The predicted model showed that costs for the long COVID group were approximately £2,500 per person per year (predicted mean cost: £2,562.50, 95% CI: £2,335.60 - £2,819.22), and £1500 in the comparator group (predicted mean cost: £1,527.43, 95% CI: £1,404.33 - 1,664.45) (**Figure 3a**, average total healthcare cost). The increase in healthcare costs persisted across all healthcare types (Figure 3b-f) and as for healthcare utilisation, in those using any healthcare services, the cost ratio was highest for GPs and lowest for inpatient hospital stays. When analysing the outcomes among different healthcare sectors, we found that healthcare utilisations (**Figure 2**) and costs (**Figure 3**) were consistently higher among people with long COVID.

**Figure 3.**
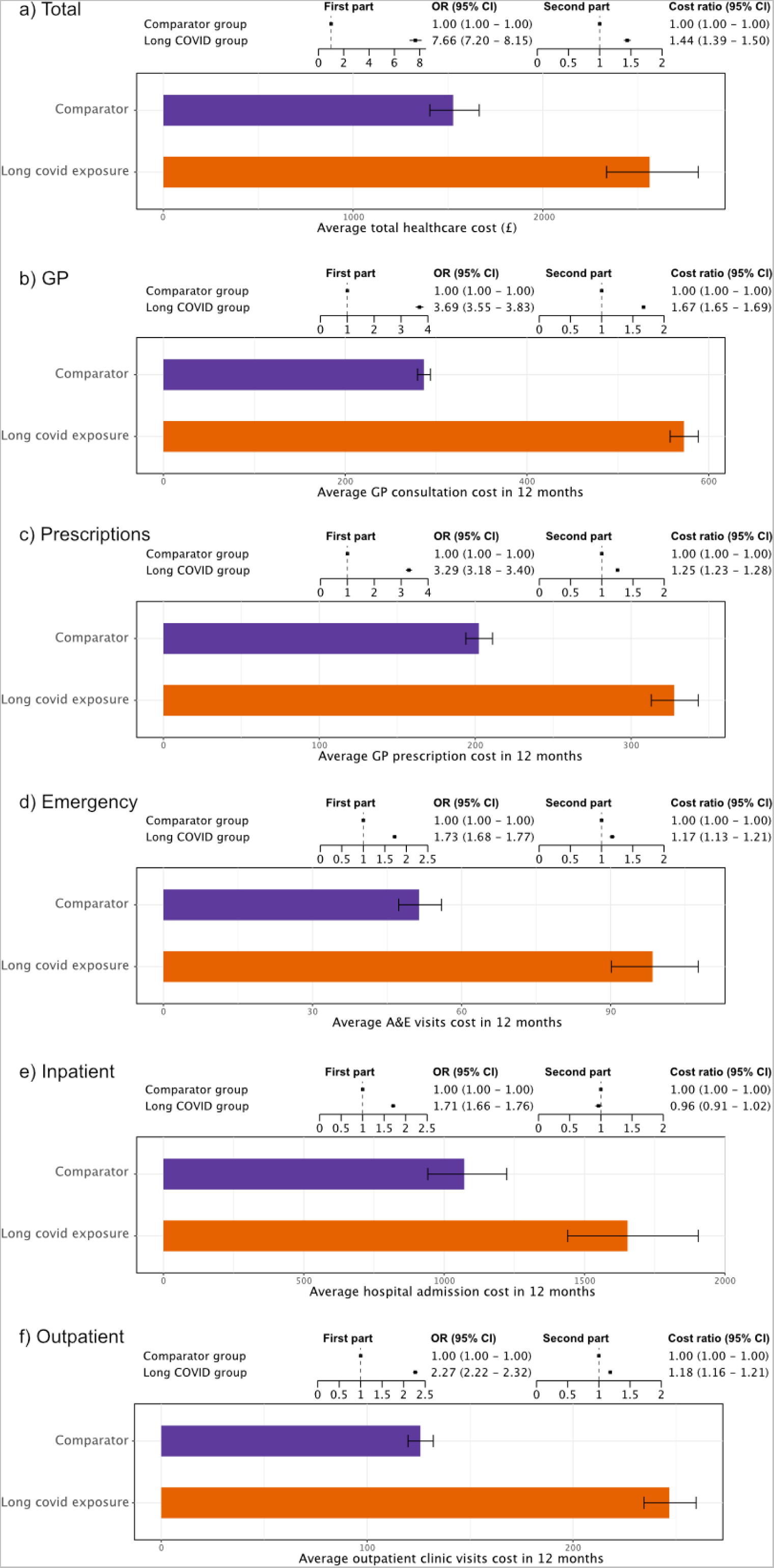
Total health costs among people with and without long COVID. Each subfigure shows the first and second part of the model in the upper panel, where the first part is the odds ratio (OR) of having any healthcare costs and the second part is the rate ratio (RR) of healthcare costs between people with long COVID and comparator groups, conditioned on people having any healthcare costs. Each bar chart shows the predicted average healthcare costs among the long COVID and Comparator groups. Values are shown for a) Total healthcare costs, and then separately for each part of the total: b) GP visits, c) prescriptions, d) Emergency care at A&E, e) Inpatient hospitalisations, f) Outpatient visits.

### Historical comparison

The difference-in-difference analyses demonstrated that individuals with a long COVID diagnosis had historically higher healthcare utilisation compared to controls, but that this difference became more marked after a long COVID diagnosis. Before the pandemic, there were approximately 20 predicted healthcare visits among the group who went on to be diagnosed with long COVID (predicted mean visits: 20.48, 95% CI:20.16 - 20.81), and 14 in the comparator group (predicted mean visits: 14.35, 95% CI:14.15 - 15.55). After the pandemic, total healthcare utilisation increased in the long COVID group to 29 (predicted mean visits: 29.28, 95% CI: 28.81 - 29.75), but remained at 14 in the comparator group (predicted mean visits: 14.05, 95% CI:13.85 - 14.24) (**Figure 4**).

**Figure 4.**
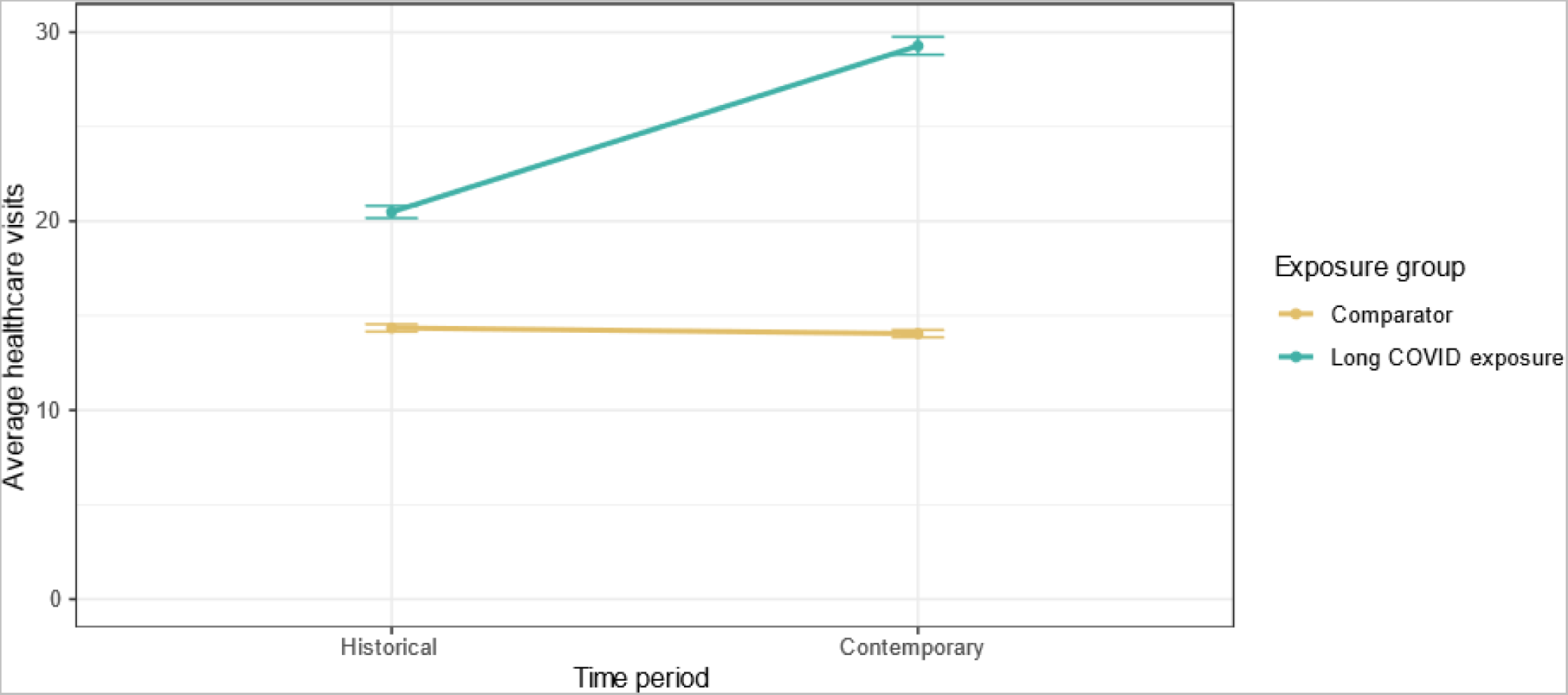
Predicted average healthcare visits before and after the pandemic. The analysis is based on a difference-in-difference analysis comparing those with long COVID and their age-, sex- and region-matched comparators.

### Factors associated with high healthcare use

In the fully-adjusted analysis model considering long COVID, we found that female sex, being obese, having asthma or mental health issues, having more comorbidities, and being previously admitted to hospital due to COVID were consistently associated with increased healthcare utilisation in both parts of the model (**Figure 5**).

**Figure 5.**
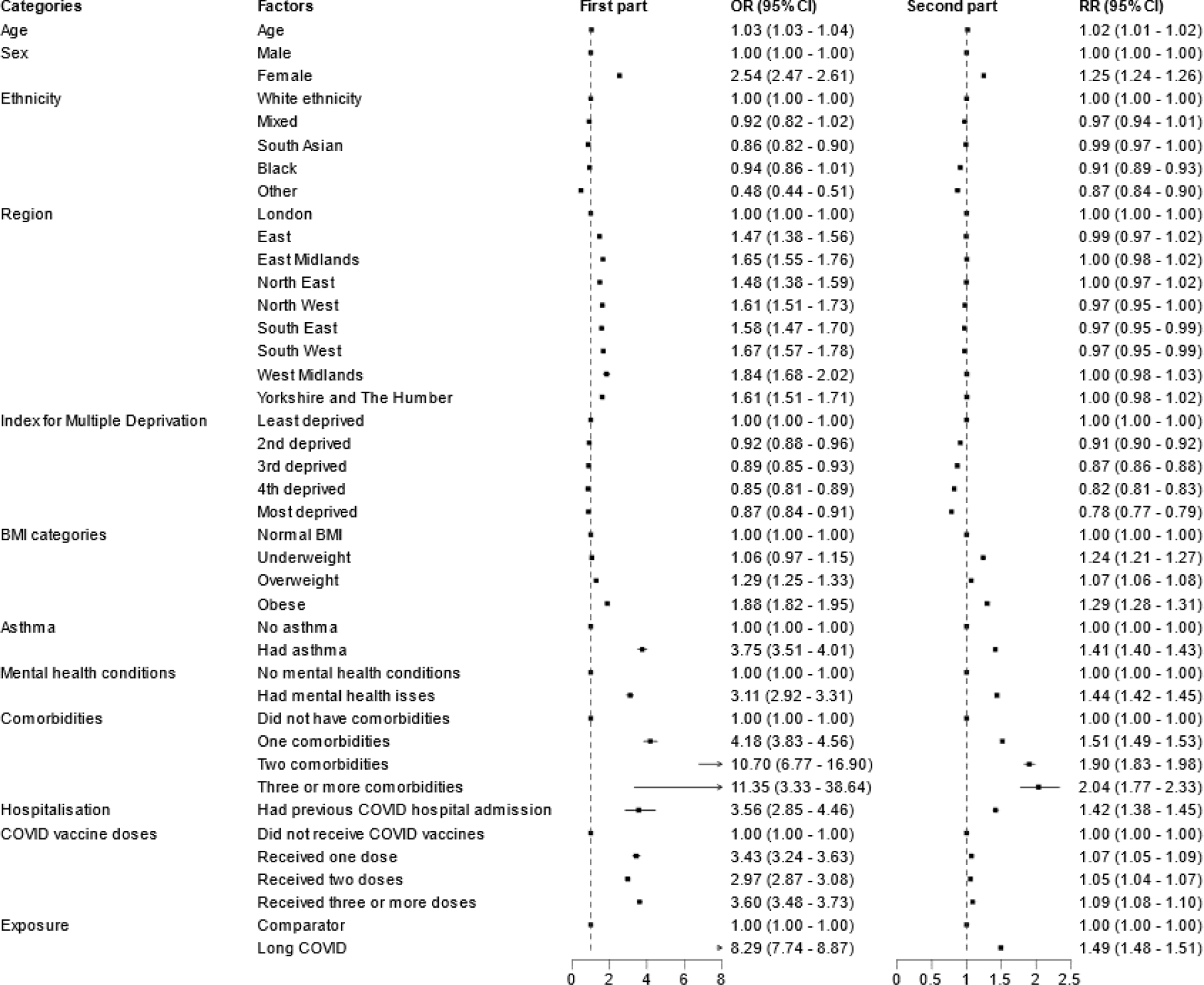
Factors associated with high healthcare use from the two-part model. The first forest plot shows the odds ratio (OR) of having non-zero healthcare use from the binomial model, the second part is the rate ratio (RR) for healthcare use from the truncated negative binomial model.

### Sensitivity analysis

After imputing missing cost data, the odds of non-zero healthcare costs and the cost ratio increased for long COVID patients, despite a slight decrease in average total cost (**Supplementary Fig 5**). In our stratified analyses, the association between long COVID and non-zero healthcare visits was more pronounced among individuals who had been hospitalised due to COVID, female sex, and those aged 40 to 69 (**Supplementary Fig 6**). However, when conditioned on non-zero healthcare resource utilisation the rate ratio was lower among the previously hospitalised stratum (**Supplementary Fig 6a**), while females and people aged 30 to 59 had a higher rate ratio (**Supplementary Fig 6b and 6c**). The predicted average healthcare visits were still higher among individuals who had previously been admitted, female sex, and older adults (**Supplementary Fig 6**.) After restricting our analyses to individuals who had been registered to a GP for one year or those who had previously tested positive, we continued to observe increased healthcare utilisations among individuals with long COVID, compared to our matched comparator group (**Supplementary Fig 7** and **Supplementary Fig 8**).

## Discussion

Our study revealed an increase in overall healthcare utilisation and associated costs in the year following a long COVID diagnosis, in comparison to those without recorded long COVID. This increase was observed across primary and secondary care including A&E visits, outpatient and inpatient stays. Our fully-adjusted models predicted that those with long COVID had nearly 30 healthcare visits per year, while the comparator group had 16 visits per year. The majority of visits in both groups were for attendance in primary care and receipt of prescriptions. The associated cost to the NHS was found to be approximately £2500 in the long COVID group, and £1500 in the comparators.

Our historical comparison demonstrated that those with long COVID were more likely to be higher users of healthcare before the pandemic compared to comparators. This indicates that those with long COVID likely had a higher pre-existing comorbidity burden than their matched comparators, as demonstrated by the differences between groups in Table 1. The change in utilisation among those diagnosed with long COVID was far greater than the change in the comparator group, indicating that long COVID may have been responsible for the increase in consultation and costs that we observed, and also highlighting the importance of adjusting for comorbidity burden in the contemporary comparison. Finally, we found that factors associated with healthcare utilisation included female sex, a history of asthma or mental health conditions, presence of comorbidities, and prior hospitalisation due to COVID-19.

Our findings relating to utilisation and costs are consistent with studies in other healthcare settings for healthcare utilisation after a COVID-19 diagnosis (4–6), and for associated healthcare costs (9). In the US, a study using Medicare data reported that among people aged over 65, people with long COVID had a higher risk of hospitalisations and outpatient visits for any cause, compared with the historical comparator group with long-term influenza symptoms (21). A study from Israel found individuals with long COVID had a higher risk of hospitalisation, home hospitalisation, and emergency visits, and an increase in costs (8), mirroring similar pre-printed findings in England (22).

A possible explanation for increased utilisation and cost is that people with long COVID attend healthcare settings separately for a variety of symptoms, for example affecting the respiratory, cardiovascular, and central nervous systems, and general non-specific symptoms. These are more likely to be reported in primary care (6,23), which could contribute to our finding of the highest primary care resource use. While effect sizes for secondary care were similar to primary care, the relatively low utilisation across A&E, outpatient and inpatient visits may relate to the nature of long COVID symptoms and the lack of effective treatment. GPs are able to offer referrals to long COVID clinics, where these are available, but any further effective treatments have so far not been identified. In addition, we also acknowledge that media coverage can influence individuals’ healthcare-seeking behaviours. For instance, a previous study on Group A Streptococcal (GAS) diseases found a correlation between extensive media coverage and increased rates of GAS testing (24). Therefore, raising awareness about long COVID may also contribute to an increase in healthcare utilisation among people with long COVID.

Our findings on factors associated with high utilisation support a recent systematic review which reported that female sex, older age, and hospitalisation, including ICU admission, are risk factors for developing post-COVID conditions (23), and are associated with high healthcare use more generally (25). In addition, in our sensitivity analyses, previous COVID-related hospitalisation, sex, and age group further modified the association between long COVID and healthcare utilisation.

Our study uses a large, representative EHR sample of individuals with clinically-recorded long COVID, and we used statistical methods for estimating healthcare utilisation and costs appropriate for zero-inflated data (16). Advancing previous analyses, we followed people with a long COVID diagnosis, and for a year post-diagnosis to ascertain the full impact of long COVID on healthcare utilisation. We did not restrict to individuals with a positive COVID-19 test (similar to other studies) because we found that only a fraction of people with long COVID diagnoses had previously tested positive in previous work (27). However, in sensitivity analyses we found that the results remained similar.

Key limitations are that the exposure and the outcomes were identified from EHR databases, which depend on people being registered and visiting their healthcare service providers. To address this, we conducted a sensitivity analysis among people who registered at least one year before the study follow-up and had at least one GP consultation record, and the results remained similar. Further, people with long COVID who do not have an EHR code could be misclassified to the comparator group. A limitation of our economic analysis is that we did not have the cost data for primary care, and some cost data for secondary care were also missing. For primary care we used the reported unit costs, as in other health economic studies (28). For secondary care, in a sensitivity analysis we imputed the missing data and the results remained similar. Further research with complete cost data in primary care and secondary care could estimate the cost more accurately.

We aimed here to examine NHS costs, but our patient advisory group suggested that people with long COVID frequently seek private healthcare, if they are able, and therefore our estimates of utilisation and cost will be underestimated, and importantly the use of private healthcare might exacerbate any socioeconomic inequalities in care. The extent of private healthcare use and wider societal costs of long COVID could not be captured in this study.

## Conclusions

Our study found that people with long COVID had increased healthcare utilisation and costs across all healthcare sectors, compared with people without long COVID. Differences varied by type of healthcare utilisation but persisted across all sensitivity analyses. We showed that people with a long COVID diagnosis typically had a higher historical healthcare burden, but that a long COVID diagnosis greatly increased their utilisation and the associated cost to the health service. Our study contributes to the growing body of evidence demonstrating the impact of long COVID, in terms of quality of life, use of healthcare, and cost. Long COVID may also affect people’s ability to participate in the workforce, with further economic consequences as well as inducing direct costs to affected individuals. Our results have implications for resource planning in future waves of infection. For example, when planning future vaccination programmes and policies for mitigating the effects of COVID, the impact of long COVID on the NHS and wider economy should be considered in addition to that of the acute COVID illness itself. Our findings imply that long COVID poses a considerable burden on attendances at all healthcare facilities and induces major healthcare costs for affected patients. Public health policies need to allocate resources for the prevention, treatment, and support of people with long COVID.

## Competing interest

none. BG is a Non-Executive Director at NHS Digital; he also receives personal income from speaking and writing for lay audiences on the misuse of science.

## Funding statement

This research was supported by the National Institute for Health and Care Research (NIHR) (OpenPROMPT: COV-LT2-0073)). In addition, this research used data assets made available as part of the Data and Connectivity National Core Study, led by Health Data Research UK in partnership with the Office for National Statistics and funded by UK Research and Innovation (grant ref MC_PC_20058). In addition, the OpenSAFELY Platform is supported by grants from the Wellcome Trust (222097/Z/20/Z); MRC (MR/V015737/1, MC_PC-20059, MR/W016729/1); NIHR (NIHR135559, COV-LT2-0073), and Health Data Research UK (HDRUK2021.000, 2021.0157).

The views expressed are those of the authors and not necessarily those of the NIHR, NHS England, UK Health Security Agency (UKHSA) or the Department of Health and Social Care. Funders had no role in the study design, collection, analysis, and interpretation of data; in the writing of the report; and in the decision to submit the article for publication.

## Data sharing statement

Other researchers can apply to access OpenSAFELY (see OpenSAFELY.org). We uploaded our analysis codes to GitHub: https://github.com/opensafely/openprompt_health_utilisation.

## Abbreviations

NHS: National Health Service

EHRs: Electronic health records

TPP: The Phoenix Partnership

GP: General practice

A&E: Acute and emergency care

OR: Odds ratio

RR: Rate ratio

## Data Availability

All analytic codes are available online at GitHub

https://github.com/opensafely/openprompt_health_utilisation

## Acknowledgement

We are very grateful for all the support received from the TPP Technical Operations team throughout this work, and for generous assistance from the information governance and database teams at NHS England and the NHS England Transformation Directorate. We express sincere gratitude to Drs. Ruth Costello, Edward Parker, and Viyaasan Mahalingasivam for reviewing our outputs, and to Dr. Jiunn Wang for suggesting difference-in-difference methods that enhanced our analyses.

## Notes

### Competing Interest Statement

The authors have declared no competing interest.

### Clinical Protocols

https://github.com/opensafely/openprompt_health_utilisation/blob/cd8ecce1e12018756375013cd3b27a25880d85a4/OpenPROMPT_longCOVID_healthcare_utilisation_protocol.pdf

### Author Declarations

London School of Hygiene & Tropical Medicine Research Ethics Committee gave ethical approval for this work(ref 28030)

