## Supplementary information for "Healthcare utilisation in people with long COVID: an OpenSAFELY cohort study"

### Supplementary Table 1. Long COVID SNOMED codes

| SNOMED codes | Term |
|---|---|
| 1325161000000100 | Post-COVID-19 syndrome |
| 1325181000000100 | Ongoing symptomatic disease caused by severe acute respiratory syndrome coronavirus 2 |
| 1325021000000100 | Signposting to Your COVID Recovery |
| 1325031000000100 | Referral to post-COVID assessment clinic |
| 1325041000000100 | Referral to Your COVID Recovery rehabilitation platform |
| 1325051000000100 | Newcastle post-COVID syndrome Follow-up Screening Questionnaire |
| 1325061000000100 | Assessment using Newcastle post-COVID syndrome Follow-up Screening Questionnaire |
| 1325071000000100 | COVID-19 Yorkshire Rehabilitation Screening tool |
| 1325081000000100 | Assessment using COVID-19 Yorkshire Rehabilitation Screening tool |
| 1325091000000100 | Post-COVID-19 Functional Status Scale patient self-report |
| 1325101000000100 | Assessment using Post-COVID-19 Functional Status Scale patient self-report |
| 1325121000000100 | Post-COVID-19 Functional Status Scale patient self-report final scale grade |
| 1325131000000100 | Post-COVID-19 Functional Status Scale structured interview final scale grade |
| 1325141000000100 | Assessment using Post-COVID-19 Functional Status Scale structured interview |
| 1325151000000100 | Post-COVID-19 Functional Status Scale structured interview |

**Supplementary Table 2. List of comorbidities**

| **Disease** |
|---|
| Non-haematological cancer |
| Haematological cancer[1] |
| Chronic respiratory disease |
| Chronic cardiac disease |
| Chronic liver disease |
| Stroke or dementia |
| Other neurological conditions[2] |
| Organ transplant |
| Rheumatoid arthritis |
| Systemic lupus erythematosus |
| Psoriasis |
| Other immunosuppressive conditions[3] |

1. Having haematological cancers six months before the index date; 2. Such as Huntington's disease, multiple sclerosis, motor neuron diseases, and other neurological diseases; 3. Including other permanent and temporary immunosuppressive diseases.

**Supplementary Table 3. The RECORD checklist.**

| | Item No. | STROBE items | Location in manuscript where items are reported | RECORD items | Location in manuscript where items are reported |
|---|---|---|---|---|---|
| **Title and abstract** | | | | | |
| | 1 | (a) Indicate the study's design with a commonly used term in the title or the abstract (b) Provide in the abstract an informative and balanced summary of what was done and what was found | Title page and page 2 | RECORD 1.1: The type of data used should be specified in the title or abstract. When possible, the name of the databases used should be included.<br><br>RECORD 1.2: If applicable, the geographic region and timeframe within which the study took place should be reported in the title or abstract.<br><br>RECORD 1.3: If linkage between databases was conducted for the study, this should be clearly stated in the title or abstract. | Title page and page 2 |
| **Introduction** | | | | | |
| Background rationale | 2 | Explain the scientific background and rationale for the investigation being reported | Page 4 | | |
| Objectives | 3 | State specific objectives, including any prespecified hypotheses | Page 4 | | |
| **Methods** | | | | | |

| | | | | | |
|---|---|---|---|---|---|
| Study Design | 4 | Present key elements of study design early in the paper | Page 5 | | |
| Setting | 5 | Describe the setting, locations, and relevant dates, including periods of recruitment, exposure, follow-up, and data collection | Page 5 | | |
| Participants | 6 | *(a) Cohort study* - Give the eligibility criteria, and the sources and methods of selection of participants. Describe methods of follow-up<br>*Case-control study* - Give the eligibility criteria, and the sources and methods of case ascertainment and control selection. Give the rationale for the choice of cases and controls<br>*Cross-sectional study* - Give the eligibility criteria, and the sources and methods of selection of participants<br><br>*(b) Cohort study* - For matched studies, give matching criteria and number of exposed and unexposed<br>*Case-control study* - For matched studies, give matching criteria and the number of controls per case | Page 5 | RECORD 6.1: The methods of study population selection (such as codes or algorithms used to identify subjects) should be listed in detail. If this is not possible, an explanation should be provided.<br><br>RECORD 6.2: Any validation studies of the codes or algorithms used to select the population should be referenced. If validation was conducted for this study and not published elsewhere, detailed methods and results should be provided.<br><br>RECORD 6.3: If the study involved linkage of databases, consider use of a flow diagram or other graphical display to demonstrate the data linkage process, including the number of individuals with linked data at each stage. | Page 5 |
| Variables | 7 | Clearly define all outcomes, exposures, predictors, potential | Page 6 | RECORD 7.1: A complete list of codes and algorithms used to classify | Page 6; online GitHub |

| | | | | |
|---|---|---|---|---|
| | | confounders, and effect modifiers. Give diagnostic criteria, if applicable. | | exposures, outcomes, confounders, and effect modifiers should be provided. If these cannot be reported, an explanation should be provided. |
| Data sources/ measurement | 8 | For each variable of interest, give sources of data and details of methods of assessment (measurement).<br>Describe comparability of assessment methods if there is more than one group | Page4 | |
| Bias | 9 | Describe any efforts to address potential sources of bias | Page 5 | |
| Study size | 10 | Explain how the study size was arrived at | Page 5 | |
| Quantitative variables | 11 | Explain how quantitative variables were handled in the analyses. If applicable, describe which groupings were chosen, and why | Page 7 | |
| Statistical methods | 12 | (a) Describe all statistical methods, including those used to control for confounding<br>(b) Describe any methods used to examine subgroups and interactions<br>(c) Explain how missing data were addressed<br>(d) *Cohort study* - If applicable, explain how loss to follow-up was addressed | Page 7–8 | |

| | | | | | |
|---|---|---|---|---|---|
| | | *Case-control study* - If applicable, explain how matching of cases and controls was addressed<br>*Cross-sectional study* - If applicable, describe analytical methods taking account of sampling strategy<br>(e) Describe any sensitivity analyses | | | |
| Data access and cleaning methods | | .. | | RECORD 12.1: Authors should describe the extent to which the investigators had access to the database population used to create the study population.<br><br>RECORD 12.2: Authors should provide information on the data cleaning methods used in the study. | Page 8 |
| Linkage | | .. | | RECORD 12.3: State whether the study included person-level, institutional-level, or other data linkage across two or more databases. The methods of linkage and methods of linkage quality evaluation should be provided. | Page 4 |
| **Results** | | | | | |
| Participants | 13 | (a) Report the numbers of individuals at each stage of the study (*e.g.*, numbers potentially eligible, examined for eligibility, confirmed eligible, included in | Table 10−11 | RECORD 13.1: Describe in detail the selection of the persons included in the study (*i.e.*, study population selection) including filtering based on data quality, data availability and linkage. | Supplementary Fig 3 |

| | | | | |
|---|---|---|---|---|
| | | the study, completing follow-up, and analysed)<br>(b) Give reasons for non-participation at each stage.<br>(c) Consider use of a flow diagram | | The selection of included persons can be described in the text and/or by means of the study flow diagram. |
| Descriptive data | 14 | (a) Give characteristics of study participants (*e.g.*, demographic, clinical, social) and information on exposures and potential confounders<br>(b) Indicate the number of participants with missing data for each variable of interest<br>(c) *Cohort study* - summarise follow-up time (*e.g.*, average and total amount) | Page 9 | |
| Outcome data | 15 | *Cohort study* - Report numbers of outcome events or summary measures over time<br>*Case-control study* - Report numbers in each exposure category, or summary measures of exposure<br>*Cross-sectional study* - Report numbers of outcome events or summary measures | Supplementary Table 5 | |
| Main results | 16 | (a) Give unadjusted estimates and, if applicable, confounder-adjusted estimates and their precision (e.g., 95% confidence interval). Make clear which | Page 14−16 | |

| | | | | | |
|---|---|---|---|---|---|
| | | confounders were adjusted for and why they were included<br>(b) Report category boundaries when continuous variables were categorized<br>(c) If relevant, consider translating estimates of relative risk into absolute risk for a meaningful time period | | | |
| Other analyses | 17 | Report other analyses done—e.g., analyses of subgroups and interactions, and sensitivity analyses | Page 17−18 | | |
| **Discussion** | | | | | |
| Key results | 18 | Summarise key results with reference to study objectives | Page 18 | | |
| Limitations | 19 | Discuss limitations of the study, taking into account sources of potential bias or imprecision. Discuss both direction and magnitude of any potential bias | Page 19 | RECORD 19.1: Discuss the implications of using data that were not created or collected to answer the specific research question(s). Include discussion of misclassification bias, unmeasured confounding, missing data, and changing eligibility over time, as they pertain to the study being reported. | Page 19 |
| Interpretation | 20 | Give a cautious overall interpretation of results considering objectives, limitations, multiplicity of analyses, results from similar studies, and other relevant evidence | Page 20 | | |

| | | | | | |
|---|---|---|---|---|---|
| Generalisability | 21 | Discuss the generalisability (external validity) of the study results | | | |
| **Other Information** | | | | | |
| Funding | 22 | Give the source of funding and the role of the funders for the present study and, if applicable, for the original study on which the present article is based | Title page | | |
| Accessibility of protocol, raw data, and programming code | | .. | | RECORD 22.1: Authors should provide information on how to access any supplemental information such as the study protocol, raw data, or programming code. | Page 8 |

*Reference: Benchimol EI, Smeeth L, Guttmann A, Harron K, Moher D, Petersen I, Sørensen HT, von Elm E, Langan SM, the RECORD Working Committee. The REporting of studies Conducted using Observational Routinely-collected health Data (RECORD) Statement. *PLoS Medicine* 2015; in press.

*Checklist is protected under Creative Commons Attribution (CC BY) license.

**Supplementary Table 4. Missing value across variables.**

| Variable | Total number | Missing number | Missing percentage (%) |
|---|---|---|---|
| **Age** | 317,852 | 0 | 0 |
| **Sex** | 317,852 | 0 | 0 |
| **Region** | 317,837 | 15 | 0 |
| **IMD quintile** | 312,029 | 5,823 | 1.8 |
| **Ethnicity categories** | 271,276 | 46,576 | 14.7 |
| **BMI categories** | 291,410 | 26,442 | 8.3 |
| **Long COVID diagnoses** | 317,852 | 0 | 0 |
| **COVID test positive** | 317,852 | 0 | 0 |
| **Previous COVID-19 hospitalisation** | 317,852 | 0 | 0 |
| **Mental health issues** | 317,852 | 0 | 0 |
| **Asthma** | 317,852 | 0 | 0 |
| **Number of comorbidities** | 317,852 | 0 | 0 |
| **Number of COVID vaccine received** | 317,852 | 0 | 0 |

**Supplementary Table 5. Distribution of outcome variables.**

| Outcome type | Month | Total number | Missing number | Long covid exposure | Comparator | p-value |
|---|---|---|---|---|---|---|
| **Mean total healthcare visits** | 1 | 317,852 (100.0) | 0 | 3.2 (2.9) | 1.3 (2.0) | <0.001 |
| | 2 | 317,852 (100.0) | 0 | 2.3 (2.6) | 1.2 (1.9) | <0.001 |
| | 3 | 317,852 (100.0) | 0 | 2.2 (2.5) | 1.2 (1.9) | <0.001 |
| | 4 | 317,852 (100.0) | 0 | 2.1 (2.5) | 1.2 (1.9) | <0.001 |
| | 5 | 317,852 (100.0) | 0 | 2.0 (2.5) | 1.1 (1.9) | <0.001 |
| | 6 | 317,852 (100.0) | 0 | 1.9 (2.4) | 1.1 (1.8) | <0.001 |
| | 7 | 317,852 (100.0) | 0 | 1.8 (2.4) | 1.1 (1.8) | <0.001 |
| | 8 | 317,852 (100.0) | 0 | 1.8 (2.5) | 1.0 (1.8) | <0.001 |
| | 9 | 317,852 (100.0) | 0 | 1.6 (2.3) | 1.0 (1.8) | <0.001 |
| | 10 | 317,852 (100.0) | 0 | 1.5 (2.3) | 0.9 (1.7) | <0.001 |
| | 11 | 317,852 (100.0) | 0 | 1.5 (2.3) | 0.9 (1.7) | <0.001 |
| | 12 | 317,852 (100.0) | 0 | 1.3 (2.1) | 0.8 (1.6) | <0.001 |
| **Mean GP consultations** | 1 | 317,852 (100.0) | 0 | 1.7 (1.7) | 0.5 (1.0) | <0.001 |
| | 2 | 317,852 (100.0) | 0 | 1.0 (1.4) | 0.5 (1.0) | <0.001 |
| | 3 | 317,852 (100.0) | 0 | 0.9 (1.3) | 0.5 (1.0) | <0.001 |
| | 4 | 317,852 (100.0) | 0 | 0.8 (1.3) | 0.5 (1.0) | <0.001 |
| | 5 | 317,852 (100.0) | 0 | 0.8 (1.3) | 0.4 (1.0) | <0.001 |
| | 6 | 317,852 (100.0) | 0 | 0.7 (1.2) | 0.4 (0.9) | <0.001 |
| | 7 | 317,852 (100.0) | 0 | 0.7 (1.2) | 0.4 (0.9) | <0.001 |
| | 8 | 317,852 (100.0) | 0 | 0.7 (1.2) | 0.4 (0.9) | <0.001 |
| | 9 | 317,852 (100.0) | 0 | 0.6 (1.2) | 0.4 (0.9) | <0.001 |
| | 10 | 317,852 (100.0) | 0 | 0.6 (1.2) | 0.4 (0.9) | <0.001 |
| | 11 | 317,852 (100.0) | 0 | 0.6 (1.1) | 0.3 (0.9) | <0.001 |
| | 12 | 317,852 (100.0) | 0 | 0.5 (1.1) | 0.3 (0.8) | <0.001 |
| **Mean prescription visits** | 1 | 317,852 (100.0) | 0 | 1.1 (1.3) | 0.6 (0.9) | <0.001 |
| | 2 | 317,852 (100.0) | 0 | 1.0 (1.2) | 0.6 (0.9) | <0.001 |
| | 3 | 317,852 (100.0) | 0 | 0.9 (1.2) | 0.6 (0.9) | <0.001 |

| | | | | | | |
|---|---|---|---|---|---|---|
| | 4 | 317,852 (100.0) | 0 | 0.9 (1.2) | 0.6 (0.9) | <0.001 |
| | 5 | 317,852 (100.0) | 0 | 0.9 (1.2) | 0.5 (0.9) | <0.001 |
| | 6 | 317,852 (100.0) | 0 | 0.8 (1.1) | 0.5 (0.9) | <0.001 |
| | 7 | 317,852 (100.0) | 0 | 0.8 (1.1) | 0.5 (0.9) | <0.001 |
| | 8 | 317,852 (100.0) | 0 | 0.8 (1.1) | 0.5 (0.9) | <0.001 |
| | 9 | 317,852 (100.0) | 0 | 0.7 (1.1) | 0.5 (0.9) | <0.001 |
| | 10 | 317,852 (100.0) | 0 | 0.7 (1.1) | 0.4 (0.9) | <0.001 |
| | 11 | 317,852 (100.0) | 0 | 0.7 (1.1) | 0.4 (0.8) | <0.001 |
| | 12 | 317,852 (100.0) | 0 | 0.6 (1.0) | 0.4 (0.8) | <0.001 |
| **Mean hospital admission counts** | 1 | 317,852 (100.0) | 0 | 0.0 (0.1) | 0.0 (0.1) | <0.001 |
| | 2 | 317,852 (100.0) | 0 | 0.0 (0.1) | 0.0 (0.1) | <0.001 |
| | 3 | 317,852 (100.0) | 0 | 0.0 (0.1) | 0.0 (0.1) | <0.001 |
| | 4 | 317,852 (100.0) | 0 | 0.0 (0.1) | 0.0 (0.1) | <0.001 |
| | 5 | 317,852 (100.0) | 0 | 0.0 (0.1) | 0.0 (0.1) | <0.001 |
| | 6 | 317,852 (100.0) | 0 | 0.0 (0.1) | 0.0 (0.1) | <0.001 |
| | 7 | 317,852 (100.0) | 0 | 0.0 (0.1) | 0.0 (0.1) | <0.001 |
| | 8 | 317,852 (100.0) | 0 | 0.0 (0.1) | 0.0 (0.1) | <0.001 |
| | 9 | 317,852 (100.0) | 0 | 0.0 (0.1) | 0.0 (0.1) | <0.001 |
| | 10 | 317,852 (100.0) | 0 | 0.0 (0.1) | 0.0 (0.1) | <0.001 |
| | 11 | 317,852 (100.0) | 0 | 0.0 (0.1) | 0.0 (0.1) | <0.001 |
| | 12 | 317,852 (100.0) | 0 | 0.0 (0.1) | 0.0 (0.1) | <0.001 |
| **Mean A&E visits** | 1 | 317,852 (100.0) | 0 | 0.1 (0.3) | 0.0 (0.2) | <0.001 |
| | 2 | 317,852 (100.0) | 0 | 0.0 (0.2) | 0.0 (0.2) | <0.001 |
| | 3 | 317,852 (100.0) | 0 | 0.0 (0.2) | 0.0 (0.2) | <0.001 |
| | 4 | 317,852 (100.0) | 0 | 0.0 (0.2) | 0.0 (0.2) | <0.001 |
| | 5 | 317,852 (100.0) | 0 | 0.0 (0.2) | 0.0 (0.2) | <0.001 |
| | 6 | 317,852 (100.0) | 0 | 0.0 (0.2) | 0.0 (0.2) | <0.001 |
| | 7 | 317,852 (100.0) | 0 | 0.0 (0.2) | 0.0 (0.2) | <0.001 |
| | 8 | 317,852 (100.0) | 0 | 0.0 (0.2) | 0.0 (0.2) | <0.001 |

| | | | | | | |
|---|---|---|---|---|---|---|
| | 9 | 317,852 (100.0) | 0 | 0.0 (0.2) | 0.0 (0.2) | <0.001 |
| | 10 | 317,852 (100.0) | 0 | 0.0 (0.2) | 0.0 (0.1) | <0.001 |
| | 11 | 317,852 (100.0) | 0 | 0.0 (0.2) | 0.0 (0.1) | <0.001 |
| | 12 | 317,852 (100.0) | 0 | 0.0 (0.2) | 0.0 (0.1) | <0.001 |
| **Mean outpatient clinic visits** | 1 | 317,852 (100.0) | 0 | 0.3 (0.8) | 0.2 (0.6) | <0.001 |
| | 2 | 317,852 (100.0) | 0 | 0.3 (0.8) | 0.1 (0.6) | <0.001 |
| | 3 | 317,852 (100.0) | 0 | 0.3 (0.8) | 0.1 (0.5) | <0.001 |
| | 4 | 317,852 (100.0) | 0 | 0.3 (0.8) | 0.1 (0.6) | <0.001 |
| | 5 | 317,852 (100.0) | 0 | 0.3 (0.8) | 0.1 (0.5) | <0.001 |
| | 6 | 317,852 (100.0) | 0 | 0.3 (0.8) | 0.1 (0.5) | <0.001 |
| | 7 | 317,852 (100.0) | 0 | 0.3 (0.7) | 0.1 (0.5) | <0.001 |
| | 8 | 317,852 (100.0) | 0 | 0.3 (0.7) | 0.1 (0.5) | <0.001 |
| | 9 | 317,852 (100.0) | 0 | 0.2 (0.7) | 0.1 (0.5) | <0.001 |
| | 10 | 317,852 (100.0) | 0 | 0.2 (0.7) | 0.1 (0.5) | <0.001 |
| | 11 | 317,852 (100.0) | 0 | 0.2 (0.7) | 0.1 (0.5) | <0.001 |
| | 12 | 317,852 (100.0) | 0 | 0.2 (0.6) | 0.1 (0.4) | <0.001 |
| **Mean admission costs (£)** | 1 | 6,949 (2.2) | 310,903 | 2,577.4 (4523.6) | 3,258.2 (4802.9) | <0.001 |
| | 2 | 6,286 (2.0) | 311,566 | 2,989.7 (4677.2) | 3,250.8 (4676.5) | 0.05 |
| | 3 | 5,985 (1.9) | 311,867 | 3,011.3 (4694.7) | 3,317.4 (5108.1) | 0.04 |
| | 4 | 5,875 (1.8) | 311,977 | 3,014.6 (4159.7) | 3,427.9 (4957.0) | 0.01 |
| | 5 | 5,778 (1.8) | 312,074 | 3,194.0 (5317.6) | 3,331.4 (4959.2) | 0.38 |
| | 6 | 5,608 (1.8) | 312,244 | 2,934.0 (4675.4) | 3,510.4 (5596.4) | <0.001 |
| | 7 | 5,189 (1.6) | 312,663 | 3,016.5 (5191.6) | 3,437.0 (5191.7) | 0.01 |
| | 8 | 5,152 (1.6) | 312,700 | 2,955.6 (4278.8) | 3,387.7 (4852.3) | 0.01 |
| | 9 | 4,825 (1.5) | 313,027 | 3,212.0 (4534.4) | 3,451.1 (5153.9) | 0.15 |
| | 10 | 4,546 (1.4) | 313,306 | 3,287.4 (6008.7) | 3,255.9 (5293.1) | 0.87 |
| | 11 | 4,260 (1.3) | 313,592 | 2,993.0 (3993.4) | 3,115.6 (4485.7) | 0.43 |
| | 12 | 3,845 (1.2) | 314,007 | 2,898.7 (4348.0) | 3,236.4 (5059.9) | 0.07 |
| | 1 | 7,949 (2.5) | 309,903 | 177.3 (117.4) | 161.2 (114.4) | <0.001 |

| | | | | | | |
|---|---|---|---|---|---|---|
| **Mean A&E visit costs (£)** | 2 | 7,087 (2.2) | 310,765 | 165.8 (115.3) | 160.4 (117.1) | 0.08 |
| | 3 | 6,902 (2.2) | 310,950 | 168.9 (113.6) | 159.5 (115.0) | 0.00 |
| | 4 | 6,722 (2.1) | 311,130 | 163.3 (103.4) | 161.3 (115.9) | 0.53 |
| | 5 | 6,650 (2.1) | 311,202 | 164.5 (108.2) | 157.6 (115.2) | 0.03 |
| | 6 | 6,358 (2.0) | 311,494 | 162.3 (102.8) | 159.0 (108.8) | 0.29 |
| | 7 | 6,018 (1.9) | 311,834 | 158.3 (104.2) | 156.9 (104.3) | 0.65 |
| | 8 | 5,789 (1.8) | 312,063 | 159.6 (103.8) | 159.3 (107.0) | 0.92 |
| | 9 | 5,485 (1.7) | 312,367 | 162.0 (112.5) | 156.5 (108.2) | 0.11 |
| | 10 | 5,189 (1.6) | 312,663 | 158.1 (109.2) | 157.8 (104.7) | 0.93 |
| | 11 | 4,771 (1.5) | 313,081 | 164.9 (123.7) | 156.2 (114.8) | 0.03 |
| | 12 | 4,264 (1.3) | 313,588 | 154.4 (106.2) | 155.9 (115.5) | 0.70 |
| **Mean outpatient clinic visit costs (£)** | 1 | 31,773 (10.0) | 286,079 | 89.0 (125.4) | 97.9 (139.6) | <0.001 |
| | 2 | 30,393 (9.6) | 287,459 | 91.6 (126.7) | 94.9 (128.9) | 0.04 |
| | 3 | 29,652 (9.3) | 288,200 | 91.9 (139.3) | 96.6 (129.0) | 0.00 |
| | 4 | 29,506 (9.3) | 288,346 | 94.1 (125.9) | 97.9 (128.6) | 0.02 |
| | 5 | 28,030 (8.8) | 289,822 | 97.5 (126.3) | 97.0 (125.9) | 0.73 |
| | 6 | 26,787 (8.4) | 291,065 | 93.4 (129.6) | 97.3 (128.2) | 0.02 |
| | 7 | 26,250 (8.3) | 291,602 | 94.4 (128.0) | 98.0 (127.2) | 0.04 |
| | 8 | 25,017 (7.9) | 292,835 | 96.7 (132.9) | 97.0 (128.4) | 0.87 |
| | 9 | 23,282 (7.3) | 294,570 | 93.3 (122.5) | 96.5 (127.8) | 0.07 |
| | 10 | 21,921 (6.9) | 295,931 | 94.9 (129.6) | 98.2 (127.3) | 0.09 |
| | 11 | 20,728 (6.5) | 297,124 | 94.5 (135.8) | 97.2 (125.8) | 0.17 |
| | 12 | 18,306 (5.8) | 299,546 | 92.6 (127.3) | 97.3 (125.9) | 0.03 |
| **Mean follow-up time (days)** | 1 | 317,789 (100.0) | 63 | 29.7 (2.4) | 29.7 (2.4) | 0.52 |
| | 2 | 311,081 (97.9) | 6,771 | 29.7 (2.3) | 29.7 (2.3) | 0.25 |
| | 3 | 304,922 (95.9) | 12,930 | 29.6 (2.8) | 29.6 (2.7) | 0.70 |
| | 4 | 296,704 (93.3) | 21,148 | 29.6 (2.8) | 29.6 (2.8) | 0.49 |
| | 5 | 287,733 (90.5) | 30,119 | 29.6 (2.7) | 29.6 (2.7) | 0.50 |

| | | | | | | |
|---|---|---|---|---|---|---|
| | 6 | 279,906 (88.1) | 37,946 | 29.6 (2.9) | 29.6 (2.9) | 0.66 |
| | 7 | 271,527 (85.4) | 46,325 | 29.4 (3.3) | 29.4 (3.4) | 0.23 |
| | 8 | 260,126 (81.8) | 57,726 | 29.3 (3.6) | 29.3 (3.6) | 0.80 |
| | 9 | 248,128 (78.1) | 69,724 | 29.4 (3.1) | 29.4 (3.1) | 0.83 |
| | 10 | 237,322 (74.7) | 80,530 | 29.2 (3.9) | 29.2 (3.9) | 0.50 |
| | 11 | 222,732 (70.1) | 95,120 | 28.7 (5.0) | 28.6 (5.0) | 0.73 |
| | 12 | 202,073 (63.6) | 115,779 | 28.9 (4.4) | 28.9 (4.5) | 0.51 |

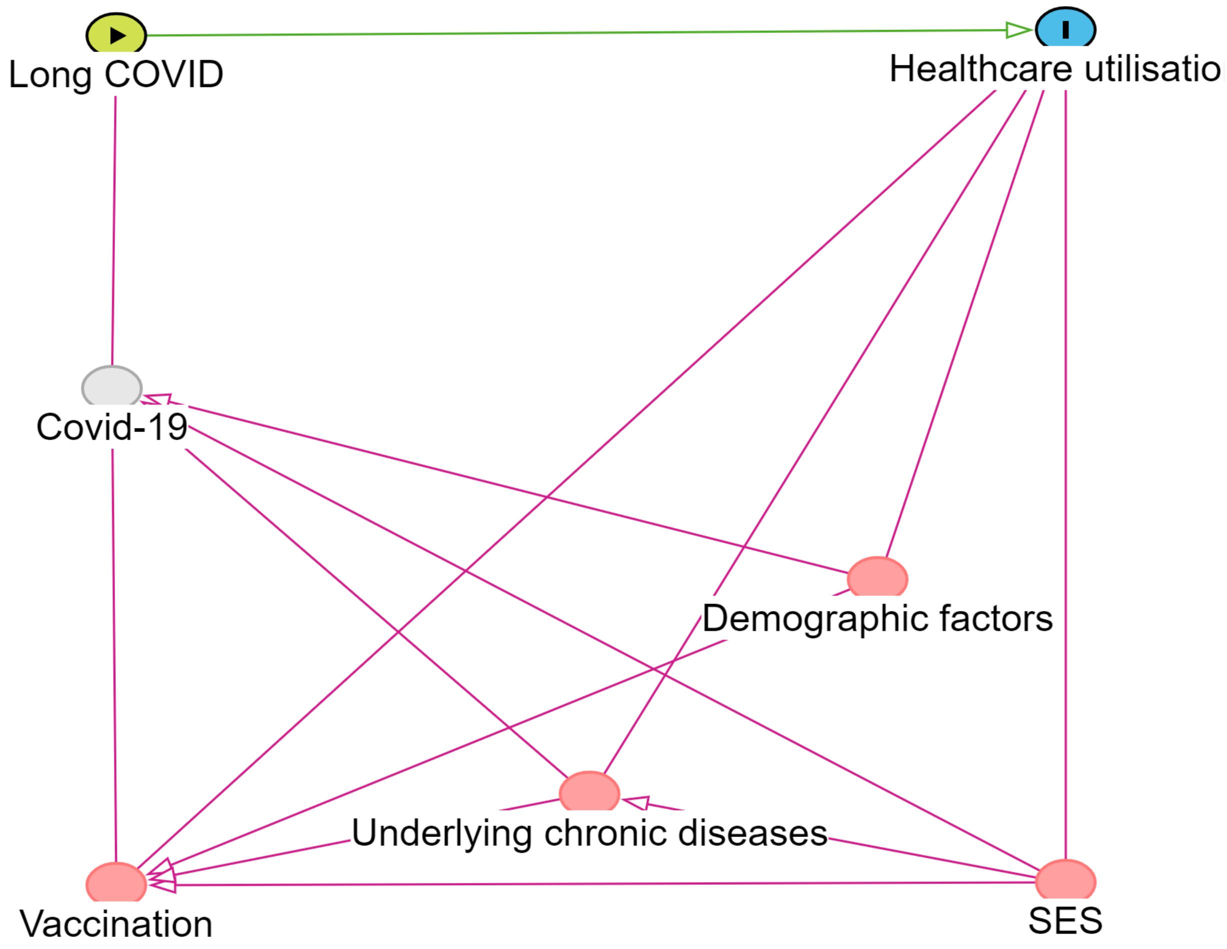

**Supplementary Figure 1.** DAGs for covariate selection.

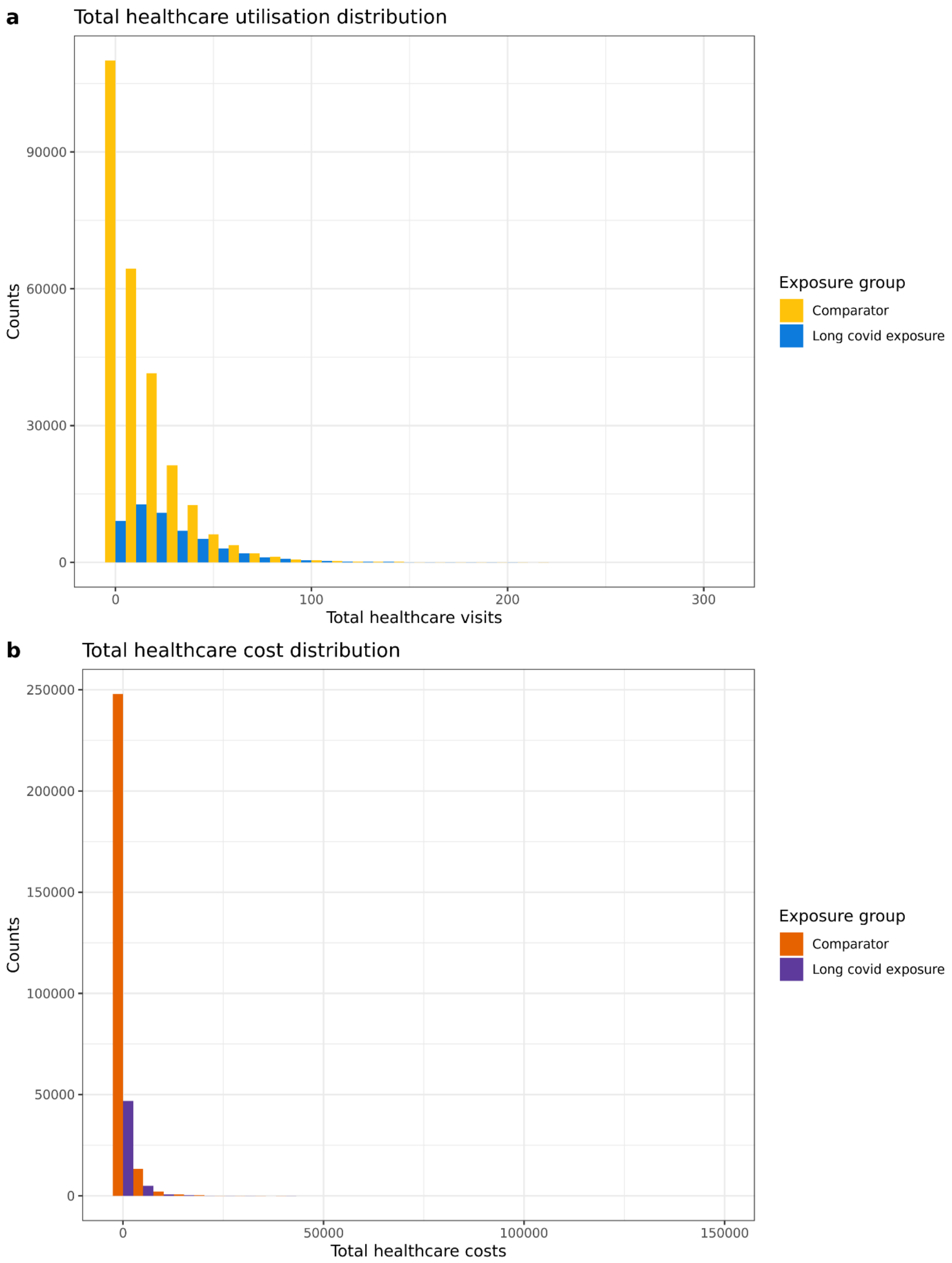

**Supplementary Figure 2. Distribution of healthcare utilisation and costs by exposure group. a. the distribution of healthcare visits; b. the distribution of healthcare costs.**

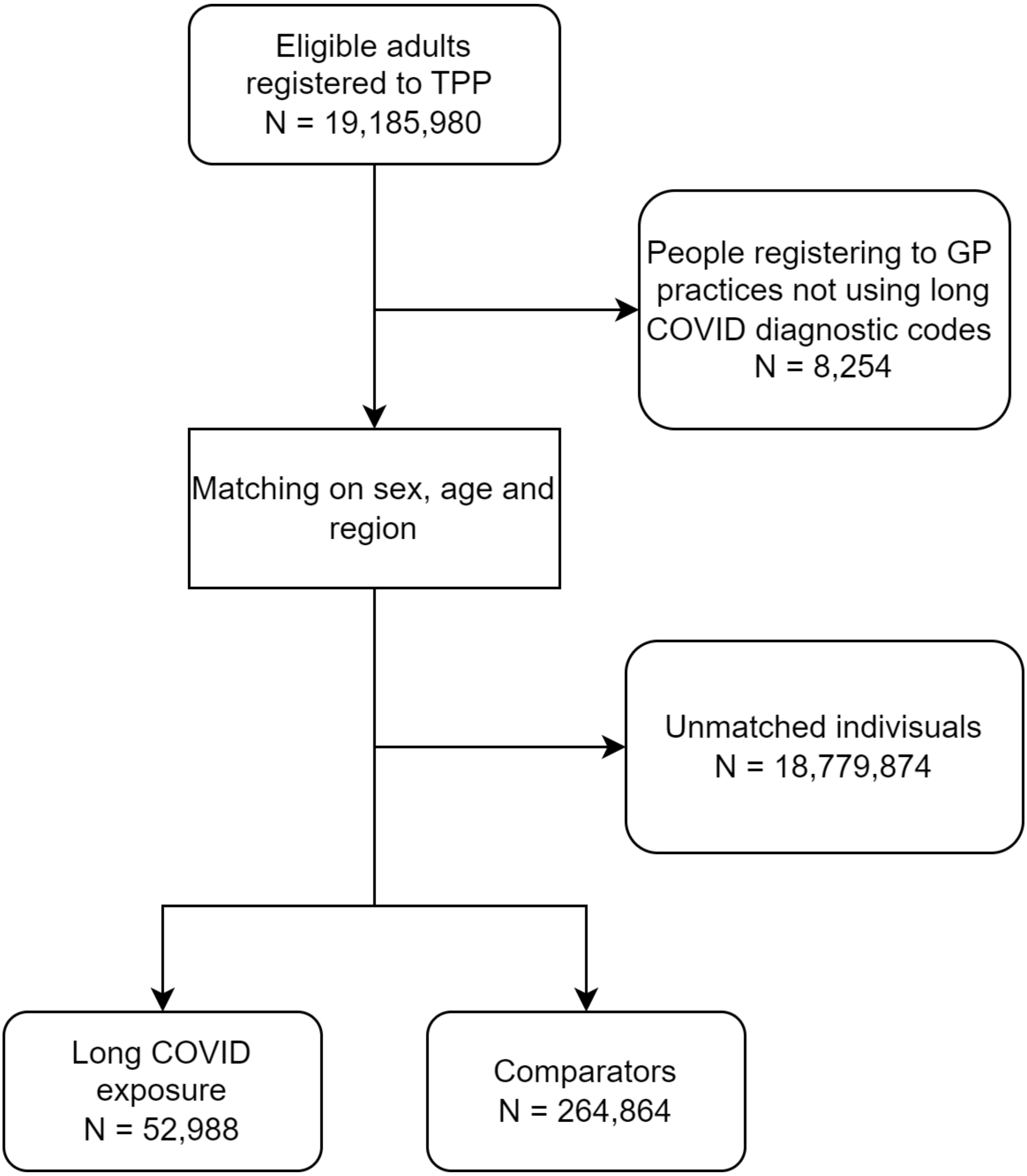

**Supplementary Figure 3. The flowchart of selecting the study population**

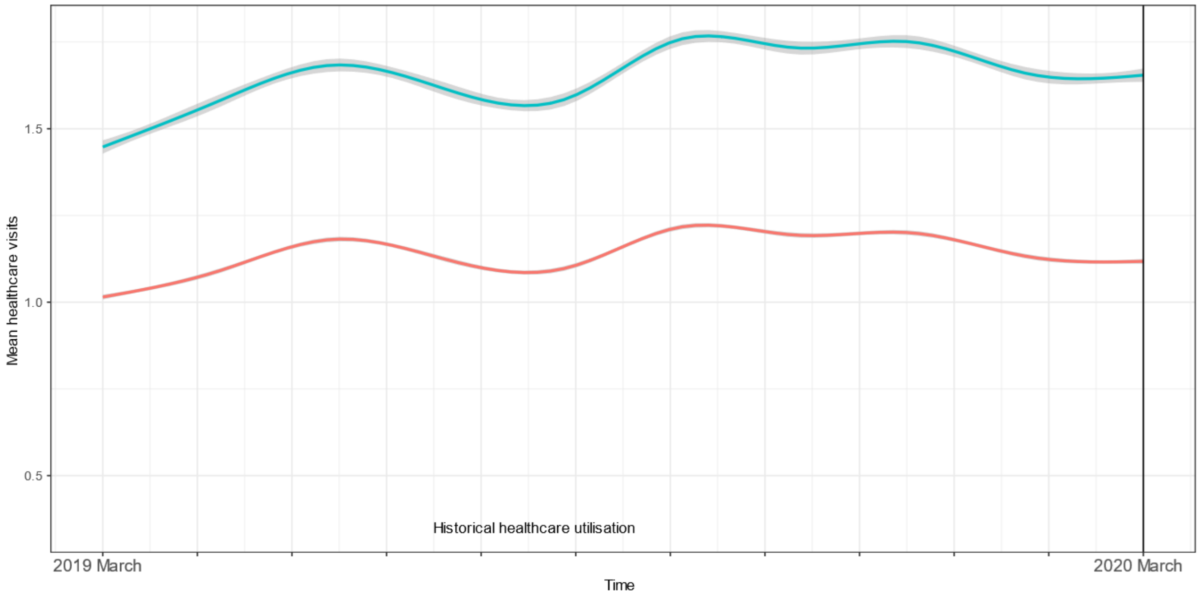

**Supplementary Figure 4. Historical average healthcare visits.** The trend in healthcare utilisation for the long COVID and comparator groups was parallel before the pandemic, meeting the common trend assumption.

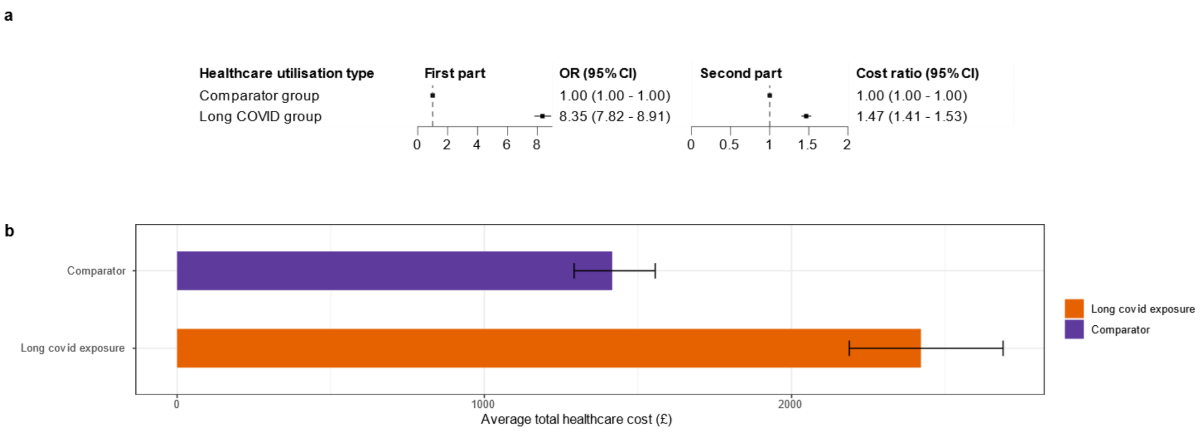

**Supplementary Figure 5. Imputed total healthcare cost**

a) Stratified by previous hospital admission due to COVID-19

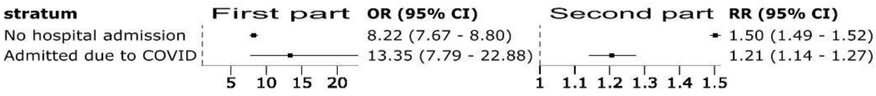

| stratum | OR (95% CI) | RR (95% CI) |
|---|---|---|
| No hospital admission | 8.22 (7.67 - 8.80) | 1.50 (1.49 - 1.52) |
| Admitted due to COVID | 13.35 (7.79 - 22.88) | 1.21 (1.14 - 1.27) |

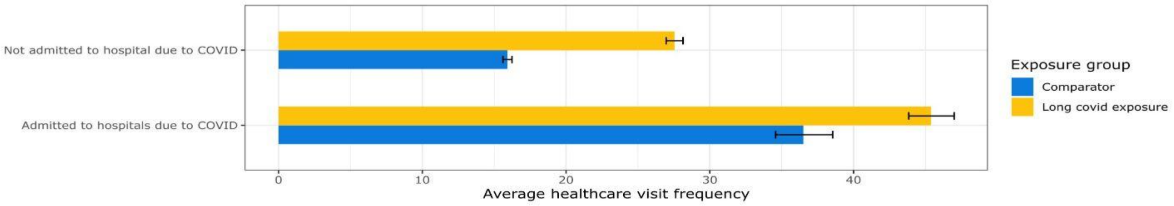

b) Stratified by sex

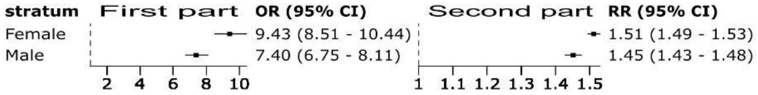

| stratum | OR (95% CI) | RR (95% CI) |
|---|---|---|
| Female | 9.43 (8.51 - 10.44) | 1.51 (1.49 - 1.53) |
| Male | 7.40 (6.75 - 8.11) | 1.45 (1.43 - 1.48) |

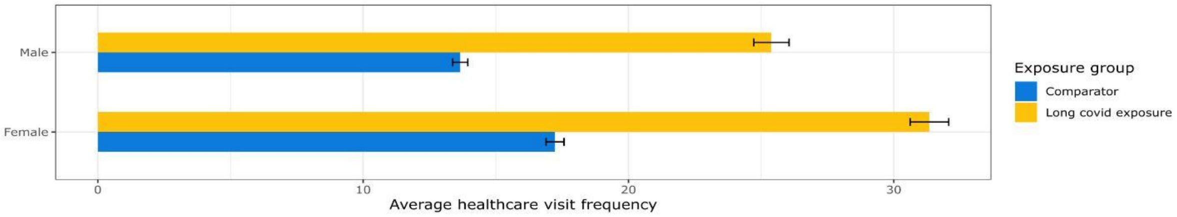

c) Stratified by age groups

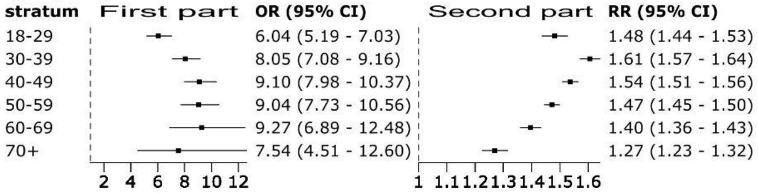

| stratum | OR (95% CI) | RR (95% CI) |
|---|---|---|
| 18-29 | 6.04 (5.19 - 7.03) | 1.48 (1.44 - 1.53) |
| 30-39 | 8.05 (7.08 - 9.16) | 1.61 (1.57 - 1.64) |
| 40-49 | 9.10 (7.98 - 10.37) | 1.54 (1.51 - 1.56) |
| 50-59 | 9.04 (7.73 - 10.56) | 1.47 (1.45 - 1.50) |
| 60-69 | 9.27 (6.89 - 12.48) | 1.40 (1.36 - 1.43) |
| 70+ | 7.54 (4.51 - 12.60) | 1.27 (1.23 - 1.32) |

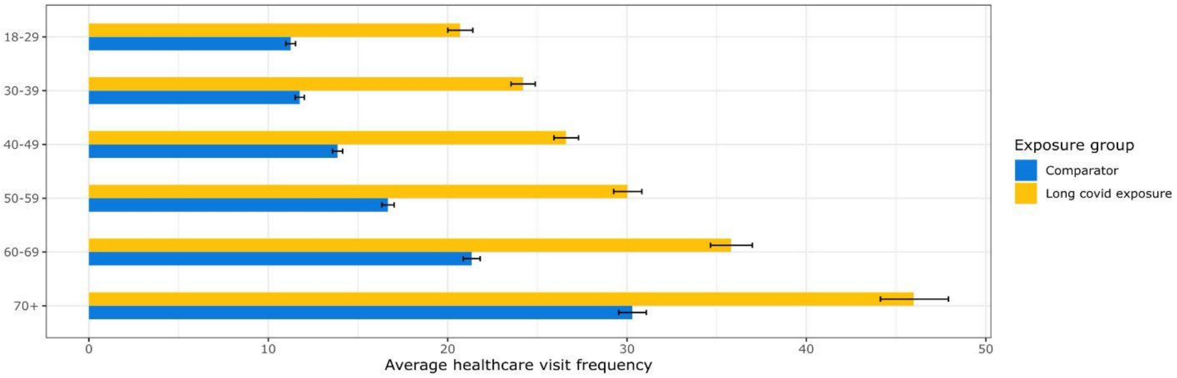

**Supplementary Figure 6. Stratified by variables**

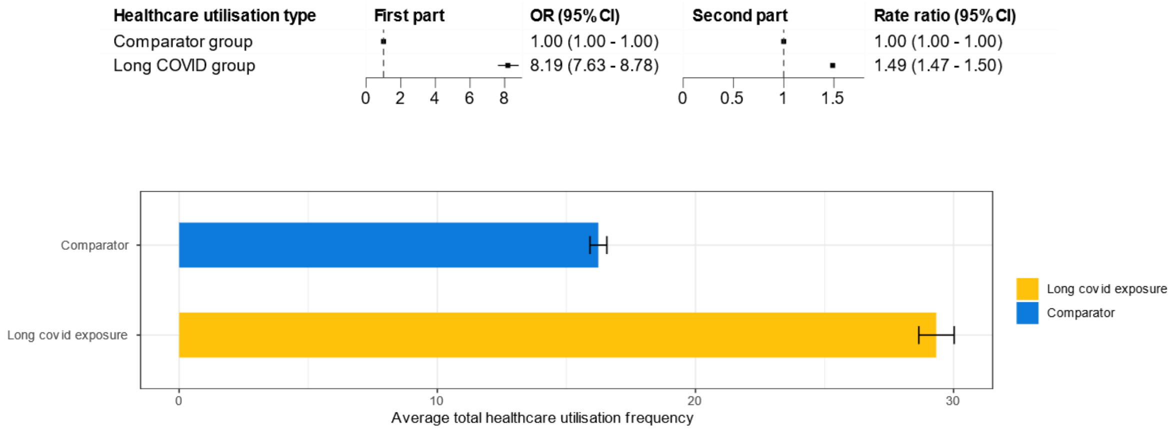

**pplementary Figure 7. Sensitivity analyses among people who had been registered a GP and visited a GP one year before the study follow-up**

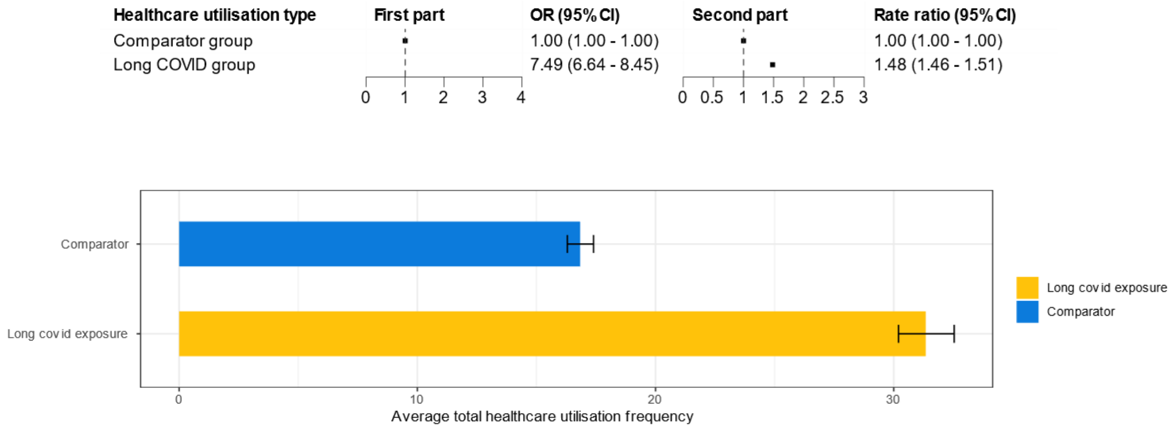

**pplementary Figure 8. Analyses among people who had tested positive for COVID fore the index date**
